# Morphological and Functional Assessment of Thyroid in Individuals with Down Syndrome

**DOI:** 10.1101/2021.05.13.21256919

**Authors:** Célia Neder Kalil Mangabeira, Rafael Kalil Mangabeira, Luis Jesuino de Oliveira Andrade

## Abstract

Individuals with Down’s syndrome (DS) present increased risk for thyroid dysfunction, especially hypothyroidism, due in increased expression of the DYRK1A gene.

**Objective:** The aim of this study was to make a morphological-functional thyroid assessment in individuals with DS.

**Materials and Methods:** This is a descriptive cross-sectional study, consisting of 29 individuals with DS, with a mean age of 12,3 ± 9,5 (0.66 – 36.00) years, 16 women (55.2%) and 13 men (44.8%), with a morphological-functional thyroid assessment being made comprising hormonal dose (Free T4, TSH), antithyroid antibody (TPOAb and TgAb) and ultrasonography of the thyroid.

**Results:** Twenty-three (79.3%) individuals presented normal thyroid function while 6 (20.7%) presented with thyroid dysfunction, 4 with hypothyroidism and 2 with hyperthyroidism. Autoimmune thyroiditis and goiter were present in 27.6% of the individuals.

**Conclusion:** Thyroid function should be assessed periodically in individuals with DS, in view of the high prevalence of thyroid dysfunction, especially autoimmune thyroiditis with consequent hypothyroidism.

## Introduction

Thyroid dysfunction in Down’s syndrome (DS) has been studied by various researchers, and a very broad range of prevalence has been estimated (1.40 to 60.34%),^1,2^ being related to increased expression of the DYRK1A gene, which can act on the thyroid gland in individuals with trisomy of chromosome 21.^3^ Among these dysfunctions, hypothyroidism presents a higher prevalence (60.34%)^2^ whereas hyperthyroidism has a lower prevalence (1.40%).^1^ The wide prevalence of thyroid dysfunction in DS occurs as a result of the study design, the sample number, age of the studied group and the geographical region studied.^4^

Alterations of the thyroid hypothalamus-hypophysis axis in DS may be associated with the primary disease of the thyroid, or the alteration of TSH secretion, dependent on a deficient dopaminergic control of hypophysary secretion.^5^

Hypothyroidism in DS, however, occurs more frequently at a more advanced age, and secondary to autoimmune thyroid disease, such as Hashimoto’s thyroiditis.^6^ Furthermore, there is subclinical hypothyroidism, which is a difficult condition to diagnose, in view of the very inconsistent clinical condition, as the signs and symptoms of this illness are confused with the signs and symptoms of DS itself, thus diagnosis fundamentally depends on hormonal assessment. In all cases of hypothyroidism, the treatment is replacement of physiological doses of levothyroxin with periodic clinical and laboratory controls. At present there is consensus about the need for periodic investigation of thyroid function in DS.^7,8^ Subclinical and transitory hypothyroidism appear to be the most frequent, and laboratory alterations could be associated with low doses of seric zinc.^9^

Hyperthyroidism in DS presents a prevalence that ranges from 1.4 to 5%, with Graves’ disease being the commonest aggravation^1^, and in the majority of cases, Hashimoto’s thyroiditis is associated in its initial stage.^10^ It is commoner to detect hyperthyroidism in DS during the period of adolescence, the youngest case having been described in an 11-month-old child with DS, and associated with diabetes mellitus type 1.^11^

The antithyroid antibodies are generated by the immunologic system of affected patients against the specific thyroid proteins that worked as antigens in these individuals. The antibodies are associated with autoimmune thyroid diseases, Hashimoto’s thyroiditis and Graves’ disease and their variations.^12^ In DS the prevalence of antithyroid antibodies is 39%, being higher in individuals with hypothyroidism (24%) and this prevalence increases with age.^13^

The ultrasonography finding of thyroid in DS is more commonly found in goiter, however, other findings, such as textural alteration as a result of lymphocytic infiltrates of Hashimoto’s thyroiditis and glandular hypoplasia is also common.^14^

The aim of the present study was to make a morphological and functional assessment of the thyroid gland in individuals with DS, participants of the “Núcleo Aprendendo Down” (Down’s learning nucleus) of the State University of Santa Cruz (UESC) of the State of Bahia, Brazil.

## Materials and Methods

This concerns a descriptive cross sectional study, conducted between 2006 and 2007 in individuals with DS, confirmed clinically, or by cariotype, participants of the “Núcleo Aprendendo Down” (Down’s learning nucleus) at UESC. The sample consisted of 29 individuals with a mean age of 12,3 ± 9,5 (0.7 – 36.0) years, 16 women (55.2%) and 13 men (44.8%), with a morphological-functional thyroid assessment being made comprising hormonal dose (Free T4, TSH), antiperoxidase antibody (TPOAb) antithyroglobulin antibody (TgAb) and ultrasonography of the thyroid.

TSH and free T4 were assessed by the chemoilluminescence method, with IMMULITE equipment, having 0.27 to 4.20 μU/mL for TSH and 0.93 to 1.7 ng/dL as reference values.

TPOAb was analyzed by electro chemoilluminescence immunoassay having up to 34 UI/mL as reference value, while TgAb was analyzed automated chemoilluminescence, having up to 115UI/mL as reference value. The antithyroid antibodies were assessed with the IMMULITE analyzer.

Sonography evaluation of the thyroid was performed using Siemens Sonolyer 60S dynamic equipment with a multifrequential linear transducer (7-10MHz), to assess topography, volume, sonographic texture and the existence of nodules.

Statistical analysis was performed using SPSS 10.0. Analysis of the associations was evaluated by the Exact Fisher and Chi-square (χ^2^) tests, at the level of significance of *P* < 0.05. The study was approved by the research ethics committee of UESC, and all the participants’ guardians signed the term of informed consent.

## Results

Twenty-three (79.3%) individuals presented normal thyroid function while 6 (20.7%) presented with thyroid dysfunction.

The prevalence of hypothyroidism was 13.8% while that of hyperthyroidism was 6.9%.

Autoimmune thyroiditis presented a prevalence of 27.6%. The antibodies TPOAb were negative in 14 individuals, and detectable in 15, being above the reference values in 8 individuals. The antibodies TgAb were negative in 14 individuals, and detectable in 15, being above the reference values in 5 individuals. Positivity of TPOAb and TgAb concomitantly and above the values of reference was observed in 5 individuals.

By ultrasonography evaluation 72.4% presented a normal exam, while 6 (20.7%) individuals presented homogeneous goiter and in 3 (6.9%) it was heterogeneous; all the individuals with goiter presented positive antibodies, however, without evidence of focal cystic or solid lesion.

In Table 1 presents a comparison of the result with reference to the association of sex, TPOAb, TgAb, and ultrasonography of the thyroid with the TSH levels.

**Table 1.** Comparison of the results with reference to the association of gender, antithyroid antibodies and ultrasonography with the TSH levels.

|  | TSH low |  | TSH high |  | TSH normal |  | Total | Statistical Test | p |
| --- | --- | --- | --- | --- | --- | --- | --- | --- | --- |
| **Sex** |  |  |  |  |  |  |  |  |  |
| Men | 0 |  | 2 |  | 11 |  | 13 |  |  |
| Women |  | 2 |  | 4 |  | 10 | 16 | Chi-square | 0.031 |
| **Total** |  | **2** |  | **6** |  | **21** | **29** |  |  |
| **TPOAb** |  |  |  |  |  |  |  |  |  |
| High | 2 |  | 4 |  | 2 |  | 8 |  |  |
| Normal |  | 0 |  | 0 |  | 21 | 21 | Fischer | 0.033 |
| **Total** |  | **2** |  | **4** |  | **23** | **29** |  |  |
| **TgAb** |  |  |  |  |  |  |  |  |  |
| High | 1 |  | 2 |  | 3 |  | 6 |  |  |
| Normal |  | 1 |  | 3 |  | 19 | 23 | Fisher | 0.027 |
| **Total** |  | **2** |  | **5** |  | **22** | **29** |  |  |
| **Ultrasound** |  |  |  |  |  |  |  |  |  |
| Normal |  | 1 |  | 4 |  | 15 | 20 |  |  |
| Abnormal |  | 1 |  | 2 |  | 6 | 9 | Fischer | 0.018 |
| **Total** |  | **2** |  | **6** |  | **21** | **29** |  |  |

As shown in the table above, there was association between gender and hypothyroidism in the sample analyzed (p 0.031). The ecographic alterations were also associated with TSH high levels (p 0.018).

When the presence of TPOAb was compared with the TSH levels, it was observed that the high levels of antibodies (>34 UI/mL) were correlated with hypothyroidism (p 0.033). Comparison between the TSH levels and the presence of TgAb also presented correlation with high levels of antibodies (p 0.027).

## Discussion

Thyroid dysfunction in DS has a high prevalence, hypothyroidism being the main complaint. In the present study, a total prevalence of thyroid alteration of 27.5% (hypothyroidism, hyperthyroidism and autoimmune thyroiditis) was found, hypothyroidism having a prevalence of 20.7% whereas the prevalence of hyperthyroidism was 6.9%. These data are compatible with those in the literature, which present a prevalence that ranges from 1.40 to 60.34%,^1,2^ however, in the present casuistic, hyperthyroidism presented a higher prevalence (6.9%) than that described in the literature (1.5 to 5%).^1^

Hypothyroidism presents a higher frequency in DS, and this can be transitory as a function of a delay in the maturation of the hypothalamus-hypophysis-thyroid axis, which leads to an increase only in the THS levels, with normal levels of free T4, characterizing subclinical hypothyroidism. Transitory elevation of TSH in individuals with DS has also been involved in low levels of seric zinc and selenium, as well as autoimmune processes.^9,15^

The occurrence of hyperthyroidism may also be present in DS, although it is less frequent, and its mechanism has not yet been well explained.^16^ In the present study, 2 individuals presented low TSH levels, and both presented presence of antithyroid antibodies.

Greater frequency of thyroid dysfunction is described in women, which tends to become equalized with the increase in age.^17-19^ The present study demonstrated that of the individuals that present alterations of the thyroid hormones, 75% were women, ranging from 5 to 36 years of age.

Autoimmune factors are related to thyroid dysfunction in DS, and the following mechanisms have been suggested: greater cellular sensitivity to interferon, the presence of the antigen HDR which would have a greater association with autoimmune e thyroiditis and the presence of enzyme 3-superoxide-dismutase-1, related to the high levels of TPOAb by the higher production of hydrogen peroxide.^20^

The present study demonstrated a high correlation between the presence of antithyroid antibodies (TPAAb and TgAb) and thyroid dysfunction, p 0.033 and 0.027 respectively. These data are demonstrated in the research by Rubello et al.^6^

Congenital hypothyroidism is present more frequently in DS (1-2%) than in the general population.^6,21-22^ The present study did not include any individuals in the age group for the detection of congenital hypothyroidism.

In the present study, once again, the high prevalence of thyroid dysfunction in DS was shown, especially hypothyroidism with autoimmune characteristics (antithyroid antibodies present), emphasizing the need for periodic monitoring of thyroid function in individuals with DS.

## Data Availability

Are available all data referred to in the manuscript.

## Notes

### Competing Interest Statement

The authors have declared no competing interest.

### Funding Statement

No external funding was received

### Author Declarations

The study was approved by the research ethics committee of Universidade Estadual de Santa Cruz - Bahia - Brazil.

## References

1. Erlichman I, Mimouni FB, Erlichman M, Schimmel MS. Thyroxine-Based Screening for Congenital Hypothyroidism in Neonates with Down Syndrome. J Pediatr. 2016; 173:165–8.

2. Amr NH. Thyroid Disorders in Subjects with Down Syndrome: An Update. Acta Biomed. 2018;89(1):132–139.

3. Thiel R, Fowkes SW. Down syndrome and thyroid dysfunction: should nutritional support be the first-line treatment? Med Hypotheses. 2007;69(4):809–15.

4. Whooten R, Schmitt J, Schwartz A. Endocrine manifestations of Down syndrome. Curr Opin Endocrinol Diabetes Obes. 2018;25(1):61–66.

5. Wilber JF, Xu AH. The thyrotropin-releasing hormone gene 1998: cloning, characterization, and transcriptional regulation in the central nervous system, heart, and testis. Wilber JF, Xu AH. Thyroid. 1998;8(10):897–901.

6. Guaraldi F, Rossetto Giaccherino R, Lanfranco F, et al. Endocrine Autoimmunity in Down’s Syndrome. Front Horm Res. 2017;48:133–146.

7. Mustacchi Z, Rozone G. A Clinica de Síndrome de Down. In _. Síndrome de Down: aspectos clínicos e odontológicos. São Paulo: CID, 1990. p. 51–98.

8. Prasher V,Gomez G. Natural history of thyroid function in adults with Down syndrome - 10-year follow-up study. J Intellect Disabil Res. 2007; 51:312–7.

9. Rasic-Milutinovic Z, Jovanovic D, Bogdanovic G, Trifunovic J, Mutic J. Potential Influence of Selenium, Copper, Zinc and Cadmium on L-Thyroxine Substitution in Patients with Hashimoto Thyroiditis and Hypothyroidism. Exp Clin Endocrinol Diabetes. 2017;125(2):79–85.

10. Bhowmick C, Grubb C. Management of multiple antibody-mediated hyperthyroidism in children with Down’s syndrome. South Med J 1997; 90:312–5.

11. Akande TO, Adeleye JO, Sepu N, Awofisoye OI. Type 1 diabetes mellitus and Graves’ disease in Down’s syndrome--a rare combination. Afr J Med Med Sci. 2016;45(3):299–301.

12. Baloch Z, Carayon P, Conte-Devolx B, et al. Laboratory medicine practice guidelines. Laboratory support for the diagnosis and monitoring of thyroid disease. Thyroid. 2003;13(1):3–126.

13. Aversa T, Salerno M, Radetti G, et al. Peculiarities of presentation and evolution over time of Hashimoto’s thyroiditis in children and adolescents with Down’s syndrome. Hormones (Athens). 2015;14(3):410–6.

14. Wu G, Zou D, Cai H, Liu Y. Ultrasonography in the diagnosis of Hashimoto’s thyroiditis. Front Biosci (Landmark Ed). 2016; 21:1006–12.

15. McGowan S, Jones J, McMillan D, et al. Screening for hypothyroidism in Down syndrome using the capillary thyroid stimulating hormone method. J Pediatr. 2015;166(4):1013-1017.e2.

16. Dos Santos TJ, Martos-Moreno GÁ, Muñoz-Calvo MT, Pozo J, Rodríguez-Artalejo F, Argente J. Clinical management of childhood hyperthyroidism with and without Down syndrome: a longitudinal study at a single center. J Pediatr Endocrinol Metab. 2018;31(7):743–750.

17. Li S, Li W, Sheng B, Zhu X. Relationship between thyroid disorders and uterine fibroids among reproductive-age women. Endocr J. 2021;68(2):211–219.

18. Al Aaraj N, Soliman AT, Itani M, Khalil A, De Sanctis V. Prevalence of thyroid dysfunctions in infants and children with Down Syndrome (DS) and the effect of thyroxine treatment on linear growth and weight gain in treated subjects versus DS subjects with normal thyroid function: a controlled study. Acta Biomed. 2019;90(8-S):36–42.

19. Iughetti L, Lucaccioni L, Fugetto F, Mason A, Predieri B. Thyroid function in Down syndrome. Expert Rev Endocrinol Metab. 2015;10(5):525–532.

20. King K, O’Gorman C, Gallagher S. Thyroid dysfunction in children with Down syndrome: a literature review. Ir J Med Sci. 2014;183(1):1–6.

21. Erlichman I, Mimouni FB, Erlichman M, Schimmel MS. Thyroxine-Based Screening for Congenital Hypothyroidism in Neonates with Down Syndrome. J Pediatr. 2016; 173:165–8.

22. Hermanns P, Shepherd S, Mansor M, Schulga J, Jones J, Donaldson M, Pohlenz J. A new mutation in the promoter region of the PAX8 gene causes true congenital hypothyroidism with thyroid hypoplasia in a girl with Down’s syndrome. Thyroid. 2014;24(6):939–44.

